# Analysis of Mutational Profile of Hypopharyngeal and Laryngeal Head and Neck Cancers Identifies *KMT2C* as a Potential Tumor Suppressor

**DOI:** 10.1101/2021.08.24.21262521

**Authors:** Marcin M. Machnicki, Anna Rzepakowska, Joanna I. Janowska, Monika Pepek, Alicja Krop, Katarzyna Pruszczyk, Piotr Stawinski, Malgorzata Rydzanicz, Jakub Grzybowski, Barbara Gornicka, Maciej Wnuk, Rafal Ploski, Ewa Osuch-Wojcikiewicz, Tomasz Stoklosa

## Abstract

Hypopharyngeal cancer represents one of worst types of head and neck tumors with only few studies focusing on the genomic profile of this type of cancer. Using targeted next-generation sequencing we analyzed 48 HPV-negative tumor samples including 23 originating from hypopharynx, and 25 from larynx. Among genes previously described as significantly mutated, *TP53, FAT1, NOTCH1, KMT2C* and *CDKN2A* were found to be most frequently mutated. We also found that more than three-fourths of our patients harbored candidate actionable or prognostic alterations in genes belonging to RTK/ERK/PI3K, cell-cycle and DNA-damage repair pathways. Using previously published data we compared 67 hypopharyngeal cancers to 595 head and neck cancers from other sites and found no prominent differences in mutational frequency except for *CASP8* and *HRAS* genes. Since we observed relatively frequent mutations of *KTM2C* (MLL3) in our dataset, we analyzed their role in vitro by generating *KMT2C*-mutant hypopharyngeal cancer cell line FaDu with CRISPR-Cas9. We demonstrate that *KMT2C* loss-of-function results in increased colony formation and proliferation, in concordance with its previously published results. In summary, our results show that mutational profile of hypopharyngeal cancers might be similar to the one observed for other head and neck cancers with respect to minor differences and includes multiple candidate actionable and prognostic genetic alterations. We also demonstrate for the first time that *KMT2C* gene may play a role of tumor suppressor in head and neck cancer, which opens new possibilities in the search for new targeted treatment approaches.

## 1. Introduction

Head and neck squamous cell carcinoma (HNSCC) remains in the top five most common human cancers with significant morbidity and mortality worldwide despite several new treatment modalities tested in this type of cancer in the last decade. Among HNSCC, hypopharyngeal cancer was diagnosed in more than 80 000 individuals in 2018 (0.4% of all sites) and was responsible for almost 35 000 deaths (0.4% of all cancer-related deaths) [1]. Smoking, human papilloma virus (HPV) infection and alcohol consumption are among major risk factors for this type of tumor. Although hypopharyngeal cancer is rare and accounts for 2–14% malignancies in the HNSCC group [2-4], the clinical prognosis is very poor, irrespectively of aggressive multidisciplinary treatments protocols and the rates of 5-year overall and disease-specific survival range at 30-35% in contrast to the laryngeal cancer, [5-7]. Unfavorable prognosis results from large percentage (60–85%) of newly diagnosed patients with hypopharyngeal cancer in advanced stages of the disease (III–IV) [6-8]. Asymptomatic progression in early disease stages, the tendency to submucosal spread and the high amount of lymphatic vessels in hypopharyngeal mucosa contribute significantly to the increase in tumor-node-metastasis (TNM) staging system of hypopharyngeal cancer [4,9]. Currently, the accepted prognostic indicators of hypopharyngeal cancer include the clinical advancement and patient’s age. Consistently with other head and neck cancers, the squamous cell carcinoma is the most common pathological type of hypopharyngeal carcinoma [10]. Contrary to other types of head and neck cancers originating from larynx, oral cavity and oropharynx, the incidence rates of local recurrence, nodular metastasis and second primary tumors are significantly higher for hypopharyngeal cancer [4,9,11]. Even the introduction of new treatment protocols in the 1990s from primary surgical resection to definitive radiation therapy combined with induction or concurrent chemotherapy, did not appear equivalently effective in improving survival rates in hypopharyngeal cancer [12]. Moreover, the advanced stages of cancers involving both larynx and hypopharynx may still be not differentiated according to the primary site. However, with respect to the clinical course and prognosis, such attitude is not rational. Unfortunately, still no individualized therapeutic options can be recommended for hypopharyngeal cancer patients. Therefore, comprehensive analysis of genetic alterations in clinically and histopathological confirmed hypopharyngeal and laryngeal cancers may help to better understand significant differences in the molecular pathogenesis of both cancers.

Currently, the identification of the underlying molecular mechanisms involved in hypopharyngeal cancer with exploration of the differentially expressed genes, key functional pathways and molecular biomarkers gives the most promising opportunity to improve the efficacy of diagnostic and therapeutic strategies among patients. Although HNSCC genetics has been widely explored by large consortia such as TCGA, these studies have significant underrepresentation of hypopharyngeal cancer [13,14]. It has been recently updated and partially supplemented by Vossen et al., who performed elegant study focused on oropharyngeal cancer and comparison with the larynx and hypopharyngeal tumors [15]. However, the genetic landscape of this type of tumor is still only partially understood. Importantly, there is still no translation of the described genetic aberrations into better stratification of patients or what is most desired, into clinical benefit.

Therefore, we performed comprehensive analysis of the molecular landscape of hypopharyngeal cancer and laryngeal cancer to analyze the mutation frequency in major genes and functional pathways associated with oncogenesis as well as to identify clinically relevant and recurrent genetic aberrations. Additionally, we compared data from our hypopharyngeal tumors with previously published results to better characterize mutational profile of this cancer. From the notable genetic aberrations identified in our patients, we have selected *KMT2C* gene, as a potential tumor suppressor inactivated in a significant proportion of patients. The role of *KMT2C* inactivation is not characterized in this type of tumor in contrast to other top-mutated genes such as *TP53* or *NOTCH1*. We tested the potential mechanistic role of *KMT2C* gene aberrations in a FaDu hypopharyngeal cell line model by using CRISPR-Cas9-targeted gene inactivation followed by functional assays.

## 2. Materials and Methods

### 2.1. Patients and Pathologic Classification

48 samples obtained from HNSCC patients were included in the analysis (Table 1). Basing on clinical and pathological data tumors were rigorously classified as hypopharyngeal or laryngeal. Blood samples were collected from 13/48 patients prior to chemo-/radiotherapy or surgical intervention and those blood (normal)-tumor pairs were subject to exome sequencing. All of the diagnostic protocols were reassessed in order to standardize them with the 8^th^ Edition of the American Joint Committee on Cancer Staging Manual. In dubious cases, histologic slides were re-examined. As pathologic TNM classification is applicable to postoperative material, we were not able to perform staging in 6 cases in which only small biopsies were available. Histologic types were consistent with the WHO classification of carcinomas, although in 5 cases a paucity of material did not allow to distinguish between keratinizing and nonkeratinizing squamous cell carcinomas (SCC). In few cases material was not originally properly designated and the samples were no longer available (such ones are marked as not determined in the Table 1).

**Table 1.** Patients characteristics.

| Parameter | Value |
| --- | --- |
| Number of patients | 48 |
| Age [median (range)] [years] | 60 (43 – 79) |
| Sex [n, %] |  |
| Male | 45 (94) |
| Female | 3 (6) |
| Localization (primary site) [n,%] |  |
| Hypopharynx | 23 (48) |
| Larynx | 25 (52) |
| Histologic type [n,%] |  |
| SCC: |  |
| Conventional (keratinizing) | 32 (67) |
| Nonkeratinizing | 8 (17) |
| Basaloid | 3 (6) |
| Not determined | 5 (10) |
| Pathologic Stage (pTNM) [n,%] |  |
| pT1N1 | 1 (2) |
| pT2 (N0/1/2/3/x) | 8 (17) |
| pT3 (N1/2/x) | 12 (25) |
| pT4 (N1/2/3/x) | 21 (44) |
| Not determined | 6 (12) |

### 2.2. Nucleic acid Isolation

DNA was isolated either using Gentra Puregene kit (Qiagen) from fresh or snap-frozen tumor tissues and cell line pellets or with DNA Blood Mini Kit (Qiagen) from patients’ blood samples. DNA was dissolved in Buffer AE (Qiagen) and stored at +4°C. One single tumor sample was isolated from formalin-fixed paraffin-embedded tissue using E.Z.N.A. FFPE DNA Kit (Omega Bio-tek).

### 2.3. Human Papilloma Virus Testing

HPV infection status was assessed using PCR with MY11/09 primers as described before, with modifications [16]. A 20µl PCR mixture contained the following components: 1U of HotStartTaq Plus polymerase (Qiagen), 2µl 10x CoralLoad Buffer, 2×0.4µl MY09/MY11 primers (10µM), 0.4µl dNTPs (10mM), 0.8µl DNA (200ng) and 15.88 µl water. Cycling conditions were set as follows: 35 cycles (60 s at 94°C, 60 s at 55°C, 60 s at 72°C) with initial denaturation for 5 min at 95°C and final extension for 7 min at 72°C.

### 2.4. TERT Promoter Sequencing

DNA sequences of *TERT* promoter (*pTERT*) were obtained in two reactions. The PCR reaction was performed with two primers: forward 5’-CACCCGTCCTGCCCCTTCACCTT and reverse 5’-GGCTTCCCACGTGCGCAGCAGGA (10 µM) [17] and the KAPA HiFi HotStart Ready Mix (KAPA Biosystems). The reaction mixture (20 µl) contained the following components: water - 7.8 µl, HotStart Mix - 10 µl, primers (forward and reverse) - 2×0.6 µl (10µM) and DNA - 1 µl (approx. 100-300ng). PCR was performed for 32 cycles (20 s at 95°C, 30 s at 72°C) with initial denaturation for 3 min at 95°C and final extension for 1 min at 72°C. The reaction resulted in formation of 147 bp product. Subsequently, amplicon was purified using VAHTS DNA Clean Beads (Vazyme) (with a 1:1.8 DNA to beads ratio) and labeled using BigDye Terminator v3.1 Cycle Sequencing kit (Thermo Fischer Scientific) and primers used in the first reaction. The reaction mixture (10 µl) contained the following components: BigDye - 0.4 µl, Sequencing Buffer - 3.6 µl, water - 2 µl, primer (forward or reverse, 1 µM) - 2 µl and 2 µl of the purified amplicon. PCR was performed in a thermocycler for 55 cycles (10 s at 95°C, 15 s at 50°C, 90 s at 60°C) with an initial denaturation for 5 min at 95°C and a final extension for 5 min at 60°C. All PCR amplicons were analyzed on 1-2% agarose gels stained with ethidium bromide (Sigma-Aldrich).

Finally, 10 µl of labeled product was purified using beads by incubating the product for 5 min with 10 µl beads and with 42 µl of 90% ethanol at room temperature, two washes with 90% ethanol for 30 seconds on a magnet and elution with 10 µl of water. The product was then processed for sequencing on a Genetic Analyzer 3500 (Applied Biosystems). Chromatograms were analyzed using FinchTV software.

### 2.5. Next-Generation Sequencing (NGS)

The whole process of library construction and enrichment was carried using SeqCap EZ chemistry (Roche NimbleGen), according to SeqCap EZ Library SR User’s Guide v.4.2, with minor modifications, briefly described below. DNA from patient samples and FaDu cell lines was converted into DNA fragment libraries using KAPA Lib Prep kit (Kapa Biosystems). 100–1000ng of DNA was sheared on Covaris M220 for 225s (175s for partially fragmented FFPE DNA) and used as an input for library construction, followed by end-repair, adenylation and adapter ligation steps. The resulting libraries were then subject to dual-sided SPRI size-selection method and PCR amplification for 3-7 cycles, depending on DNA input. Subsequently, libraries were mixed into 8- to 24-plex pools, hybridized to SeqCap EZ capture probes and reamplified for 9–10 PCR cycles.

For custom sequence capture SeqCap EZ custom probe designs targeting 7 Mb and 10Mb were used and for exome sequencing SeqCapEZ Exome v2 or MedExome (Roche NimbleGen) probe sets were employed. Additionally, TruSeq library preparation and exome enrichment (Illumina) kits were used according to user guides for sequencing of three normal-tumor sample pairs. Regions not overlapping between custom/exome panels were excluded from analyses when needed. List of probe designs used for individual samples is provided in Supplementary data 1.2.

DNA and DNA library concentrations were measured on NanoDrop (Thermo Fisher Scientific) and Qubit fluorometer using dsDNA High Sensitivity kit (Thermo Fisher Scientific). Quality of DNA and DNA library fragment size ranges were assessed on 0.7–2% agarose gels or 2100 Bioanalyser instrument (Agilent Technologies) when needed.

All libraries were sequenced on Illumina HiSeq 1500 instrument using 2 × 100 bp reads. Tumor samples analyzed with custom panels were sequenced to reach mean coverages in range 49.2–234x (median 154.93x) with % bp @ 20x in range 77.7–97.2% (median 95.6%) while tumor exome samples were sequenced to reach mean coverages in range 31.5– 145.2x (median 112.3x) and % bp @ 20x in range 71.4–99.2% (median 92.2%). Blood samples were sequenced to reach mean coverages in range 52.6– 166.9x (median 93.4x) and % bp @ 20x in range 80.1–98.5% (median 89.8%).

### 2.6. NGS Data Acquisition and Analysis

Raw sequencing data was processed according to Broad Institute recommendations. Variant discovery included following steps: quality control of raw fastq, adapter trimming and low-quality reads removal using Trimmomatic [18], read mapping to hg19 using BWA-MEM [19], duplication removal, local re-alignment and quality recalibration using GATK and Picard and variant calling using UnifiedGenotyper, HaplotypeCaller. Exome data was additionally analyzed using Mutect-2 to identify somatic single nucleotide variants (SNV) and insertions-deletions (indels) through direct comparison of germline and tumor samples [20].

Variants were filtered using public (NHLBI ESP [21], gnomAD [22] and internal databases in order to remove common genetic variation. CADD ([23]), PolyPhen2 [24], CHASM [25], SIFT [26], FATHMM [27] and Mutation Taster [28] predictions were used to identify possible protein-damaging missense alterations. All variants were also manually curated to avoid including sequencing artifacts and ClinVar [29], Varsome [30] and COSMIC [31] databases were also used to aid final variant classification. Complete lists of classified variants are available in Supplementary data 3.

Copy-number calling was performed by sequencing coverage analysis using CNVkit v0.9.5 [32]. CNVkit was run with default settings except for 400bp bin size limit. Groups of normal samples were used to create reference coverage models across predefined targets for each custom/exome capture. Two samples were recentered due to clear deviation of basal copy number. Additionally, a GISTIC 2.0.23 [33] analysis was performed on segmented CNV data acquired for genomic regions targeted by all custom captures (Supplementary data 1.3). GISTIC was run with default parameters except for 5000bp pseudo-markers spacing setting. TCGA CNV data was reanalyzed with identical GISTIC setting. RAW CNV segmentation data for patients and cell lines is available in Supplementary data 3.2 and 3.5.

Heterozygosity alterations were analyzed across covered regions by plotting a chart of the observed absolute variant allele frequency (VAF) deviations from 0.5 (heterozygous state) for all variants called by HaplotypeCaller, with allele frequencies in range (0.0001, 0.95) in gnomAD and coverage larger than 30x. Normal-tumor pairs were additionally analyzed in a similar manner that also included calculation of VAF shifts between normal and tumor samples. This data was also segmented using CNVkit built-in cbs method after exclusion of homozygous variants to aid in identification of regions with allelic imbalance. Ambiguous results with VAF deviation lower than 0.15 across segment were considered not altered.

External mutational data for HPV-negative (HPVneg) head and neck cancers, was downloaded via cBioPortal [34,35] for TCGA PanCancer Atlas [36], Agrawal et al. [13] and Stransky et al. [14] projects or from journal’s site for Vossen et al. [15] project (referred also as “NKI dataset”). Datasets from those projects combined with current dataset referred as “Medical University of Warsaw (MUW) dataset” are further referred as “combined dataset”.

Text data was parsed using python 2.7 and pandas 0.22. Plots and statistical analyses for NGS data were generated using R library maftools v.2.3.40 [37] and matplotlib 2.2.4 [38].

### 2.7. Cell Culture

All in vitro experiments were performed on the human cell line derived from squamous cell carcinoma of the hypopharynx, namely FaDu (HTB-43; ATCC). Cells were typically cultured in 75 cm^2^ adherent cell flasks in DMEM D6429 medium (Sigma) supplemented with 10% HyClone calf serum (FBS, SH30072.03, GE Healthcare) and 1x antibiotic/antimycotic-solution (30-004-Cl, Corning) or penicillin-streptomycin solution (P4333, Sigma) during lentiviral infections. Cells were detached into suspension with 1x Trypsin (15090-046, Gibco), passaged in 2-3 days intervals and cryopreserved in liquid nitrogen in DMEM supplemented with 10% DMSO and 50% FBS. For routine passaging, cells were counted in trypan blue in Bürker chamber. For in vitro experiments, Count and Viability Kit for Muse Cell Analyser instrument (Luminex) was used for more accurate counting. All cell cultures were monitored for Mycoplasma contamination by PCR weekly.

### 2.8. Generation of CRISPR-Cas9 KMT2C-Mutant Cells and Clone Selection

CRISPR-Cas9 system was used to generate *KMT2C*-mutant (*KMT2C*^mut^) FaDu cells. pLenti CRISPR v2 plasmid (Addgene #52961, [39]) was used along with lentiviral transfection to acquire stable expression of two different sgRNAs (sg2 and sg4), targeting exons 3 and 12 of *KMT2C*, respectively, as well as a non-targeting sgRNA control (sgNTC) (Table 2). *KMT2C*-targeting sgRNAs were designed using E-CRISPR [40] and CHOPCHOP tools [41]; sg2 sequence was also previously included in the GeCKO v2 library [39]. All used sgRNA sequences were predicted to affect the curated *KMT2C* transcript variant NM_170606.3 as well as most of putative transcripts except for XM_011516454.2, XM_017012489.1 and XM_017012490.2 lacking several exons at the N-terminus. HIV-SFFV-mRFP (received courtesy of dr Els Verhoeyen, Centre International de Recherche en Infectiologie, Lyon) plasmid was used as a transfection control. Lentiviral particles were assembled in HEK293 cells grown in DMEM supplemented with 10% FBS and PenStrep, using psPAX2 (Addgene #12260) and pMD2.G (Addgene #12259) plasmids. Virus-containing medium was acquired twice after subsequent overnight incubations and each time it was filtered with 0.45µm syringe filter, concentrated by overnight centrifugation, mixed in 1:1 proportion with fresh DMEM medium and added to FaDu cells. 24h after second infection, puromycin selection was started at 2 µg/ml concentration previously measured to decrease viability of unmodified cells below 5%, with gradual decrease to 0.5 µg/ml over 1 week, resulting in a complete detachment of control HIV-SFFV-mRFP-expressing cells and acquisition of 100% or near 100% pLentiCRISPR transfected cell population as confirmed during further clonal selection.

**Table 2.**
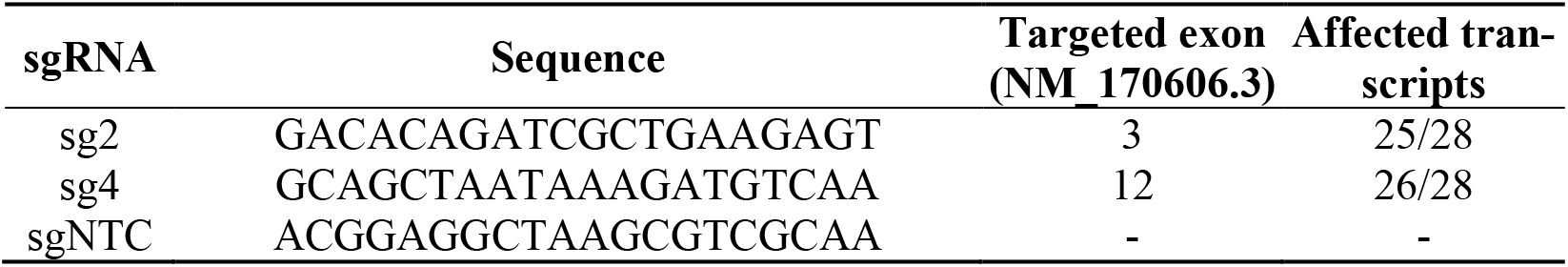
Sequences of sgRNAs used for induction of *KMT2C* mutations in FaDu cell line.

Induction of mutations by sgRNAs was confirmed by gel electrophoresis of PCR products amplified with primers flanking the sgRNA binding/Cas9 cut sites and subsequently using Sanger sequencing and NGS (Supplementary data 2.4 and data not shown).

FaDu cells expressing sg2 and sg4 were diluted to achieve an average concentration of 0.8 cells / per well when seeded on 96-well plates. Clones were detached from wells where single cells formed colonies and propagated through 24- and 6-well plates and 20 cm^2^ and 75 cm^2^ flasks when clone cell lines were frozen and DNA/RNA isolation was performed.

Clone pools and selected clones were compared to NTC-treated and non-treated cells using NGS to exclude the presence of significant off-target mutations and copy-number variations induced by CRISPR-Cas9.

### 2.9. Clonogenic Assay

Clonogenic assays for FaDu cell lines were carried out in standard 6-well plates in an amount of 625 cells per well. Cells were seeded in triplicates and cultured for approximately 14 days or until colonies started to merge, rinsed with PBS and fixed and stained with methanol and 20% crystal violet solution.

After staining, plates were scanned with BioRad GS-800 scanner. Colonies were counted with ImageJ software with Fiji package [42] using *analyze particles* function (size: 0.0001–infinity, circularity 0.01– 1.00), area covered by colonies was measured for those automatically counted.

### 2.10. DNA synthesis assay

For DNA synthesis measurements Click-iT EdU Imaging Kit (Thermo Fisher Scientific) was used and EdU incorporation measured. Cells were seeded on 6-well plates in 0.15×10^6^ cells/well density in triplicates, incubated with 10 µM EdU for 1h on the next day, detached with tripsin, fixed and stained with Alexa Fluor 488. Percentage of EdU-positive cells was measured on FACSCanto II cytometer (Becton Dickinson). Exemplary gating is provided in supplementary data 1.4.

### 2.11. Cisplatin-Sensitivity Assays

CellBlue and CellTiterGlo assays (Promega) were used to measure cell sensitivity to cisplatin. Cells were seeded in 96-well plates in 5 replicates per each group in a density of 3×10^3^ cells per well and incubated for 24h prior to cisplatin addition. Cisplatin was added to achieve 2 µg/ml and 10 µg/ml final concentrations and cells were treated for 48h. Viability measurements were taken as recommended by the manufacturer i.e. 10 minutes after addition of CellTiter-Glo and 4 hours after addition of CellTiter-Blue. Fluorescence and luminescence were measured on Victor X4 instrument (Perkin Elmer), the latter in white opaque plates. Cell-free medium was used to measure background signal.

### 2.12. Statistical analysis of in vitro data

All in vitro data was analyzed using GraphPad Prism 6. For clonogenic and DNA synthesis assays, results for KMT2C-mutant cells were compared against NTC cells using Dunnett’s test. In cisplatin-sensitivity assays, average background-subtracted readouts from cisplatin-treated cells were recalculated as fractions of their corresponding controls and compared to NTC cells using Dunnett’s test.

## 3. Results

### 3.1. Integrated Analysis of Small-Scale Mutation and Copy-Number Variation Data from laryngeal and hypopharyngeal cancers

To define the mutational profile in our cohort of patients (MUW dataset) comprised of laryngeal and hypopharyngeal cancers, we analyzed 37 genes previously described as significantly mutated in Head and Neck and Esophageal Carcinoma [14,43-49] and covered by the probe designs used to enrich genomic libraries (Supplementary data 1.1–2). Variants were manually curated to exclude benign and likely benign. Copy-number and loss-of-hetereozygosity (LOH) data was reanalyzed with the small-scale mutation data to investigate the interplay between these types of genetic aberrations.

In this approach, 32 genes were altered by small-scale mutations in our dataset (Supplementary data 2.1 and 3.1) among which 13 genes were mutated in at least 3 patients (Figure 1). The top five mutated genes were *TP53, FAT1, NOTCH1, KMT2C* and *CDKN2A. TP53* was typically biallelically altered in most cases, either by multiple mutations or combinations of mutations and shallow deletions or copy-neutral losses-of-heterozygosity (CN-LOH), most frequently involving deletion of the short arm of chromosome 17. In some cases, putative amplification of mutated allele was found. *FAT1* was altered mostly by frameshift and nonsense mutations, accompanied by shallow deletions. *NOTCH1, KMT2C* and *CDKN2A* were similarly altered by truncating mutations or/and shallow deletions or CN-LOH. Additionally, deep deletions of *CDKN2A* were detected in 20.8% of patients (10/48).

**Figure 1.**
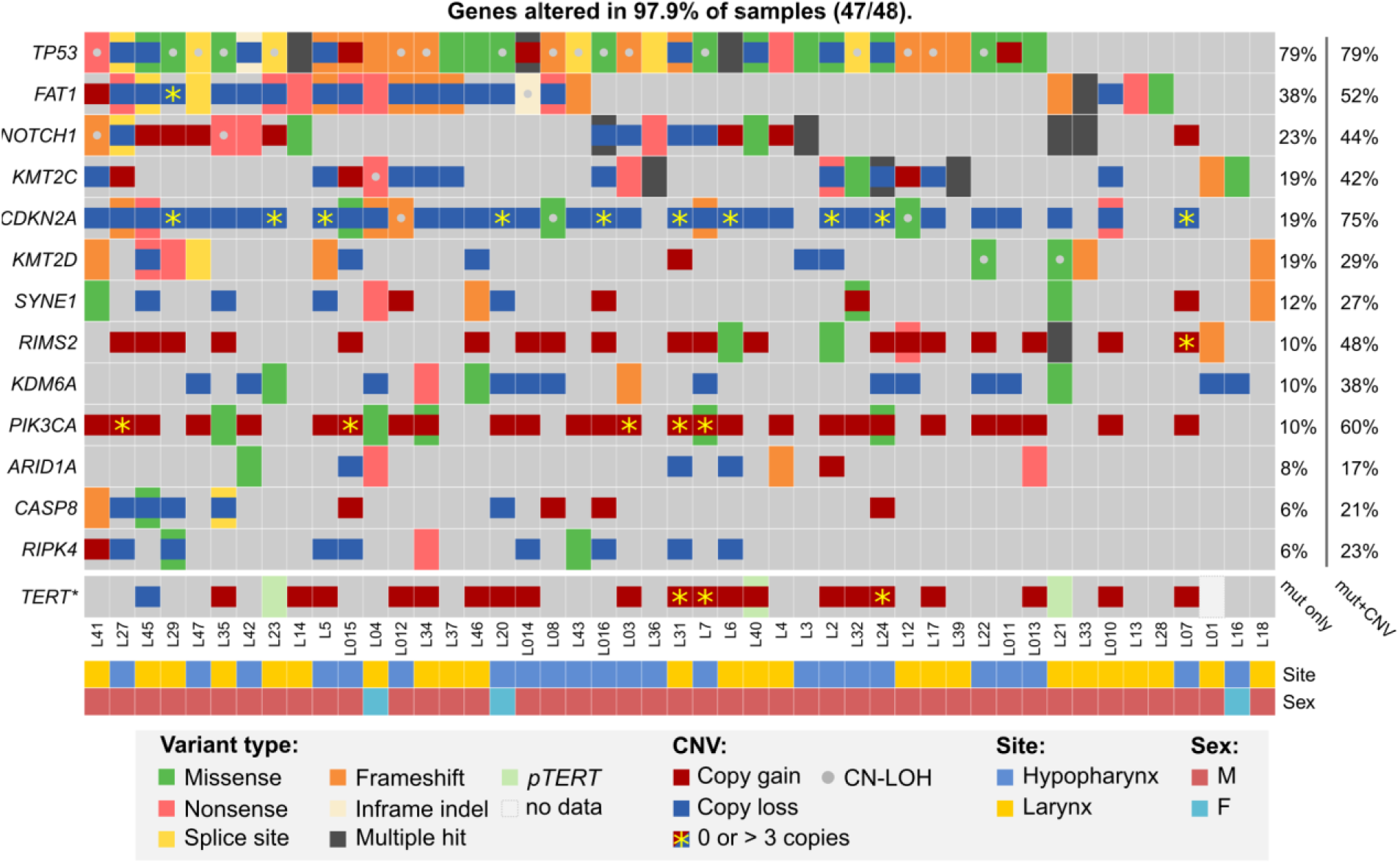
– Mutational profile of laryngeal and hypopharyngeal cancer in the MUW dataset. Selected genes previously identified as significantly mutated were analyzed for small mutations, copy-number alterations and copy-neutral duplications (CN-LOH). Only genes altered by small-scale mutations in at least 3 patients are shown. Amplifications (> 3 copies) and deep deletions (0 copies) are marked for all genes except *KDM6A* (chromosome X). Data for *TERT* (Sanger sequencing of promoter and CNV data) is presented separately at the bottom.

13 other genes were recurrently mutated in more than 2 patients (Supplementary data 2.1). Similarly to *KMT2C*, two other SET domain-containing protein encoding genes *KMT2D* and *NSD1* were affected mostly by truncating mutations or/and shallow deletions.

Since *TERT* promoter was not covered by any of our sequencing panels, we used Sanger sequencing for this analysis. Mutations of *pTERT* were detected in 6.4% (3/47) patients, for which DNA was available. None of *pTERT*-mutated patients had any significant CNV of *TERT* gene, however three other patients had *TERT* amplification (4–5 copies) (Figure 1).

A GISTIC analysis of CNV data from MUW dataset yielded a similar pattern of amplification and deletion peaks as in previously published TCGA HPVneg cohort (Supplementary data 2.2). In the significantly amplified regions, multiple genes were found to have 4 or more copies recurrently, including *CCND1* (29%, 14/48 patients), *BIRC2/3* (8.3%, 4/48), *FGFR1* (10.4%, 5/48), *TP63* (14.6%, 7/48), *EGFR* (6.3%, 3/48), *ERBB2* (4.2%, 2/48), *MDM2* (4.2%, 2/48), *TERT* (6.3%, 3/48), *PIK3CA* (5/48), *MYC* (8.3%, 4/48), *MET* (4.2%, 2/48). *ERBB2, BIRC2/3, EGFR* and *CCND1* were highly amplified in some patients (in range of 39–52, 6–16, 5–15 and 4–11 copies, respectively). In the significantly deleted regions, *CDKN2A/B* (20.8% of patients, 10/48), *FAT1* (2.1%, 1/48) and *PTEN* (2.1%, 1/48) were affected by deep deletion.

A total of 18.8% of patients (9/48) harbored *PIK3CA* mutations and/or 4-5 gene copies and 6.3% (3/48) harbored *PTEN* mutation or deep deletion. In addition to the aforementioned copy number alterations, other well-known RAS pathway-activating events were detected in 8.3% patients (4/48): hotspot mutations in *HRAS* (1 patient), *PTPN11* (1) and *FGFR3* (1) and *NF1* homozygous deletion (1). Finally, one patient carried *KRAS* amplification (5 copies).

Genes implicated in DNA repair were affected in 18.8% of patients (9/48), either by mutation and deletion or CN-LOH (*BAP1*: 1 patient; *BRCA2*: 2; *CHEK2*: 1; *PMS2*: 1) or by mutation only (*ARID1A*: 4 patients, *BRCA1*: 1; *RECQL*: 1). Additionally, 25% of patients (12/48) were found to carry shallow deletions of *ATM* also identified as a significant GISTIC deletion peak yet without any associated *ATM* mutations.

4.2% (2/48) patients carried potentially inactivating *CREBBP* mutations, similar to those previously described in leukemia [50]. Truncating mutations in *ASXL1* in 8.3% (4/48) and *TET2* in 6.3% (3/48) patients were also detected.

We then analysed NGS data for potentially druggable or prognostic genetic alterations. We selected only previously described mutations or those with strong prediction of pathogenicity, deep deletions and amplifications to at least 5 copies, while eliminating shallow deletions without a second hit, low-level amplifications and other ambiguous alterations. In such strict approach, we found that over three-fourths of patients harbored at least single alterations of genes involved in RTK/RAS/PI3K, cell-cycle or DNA damage repair pathways (Figure 2, Suplement S3.3).

**Figure 2.**
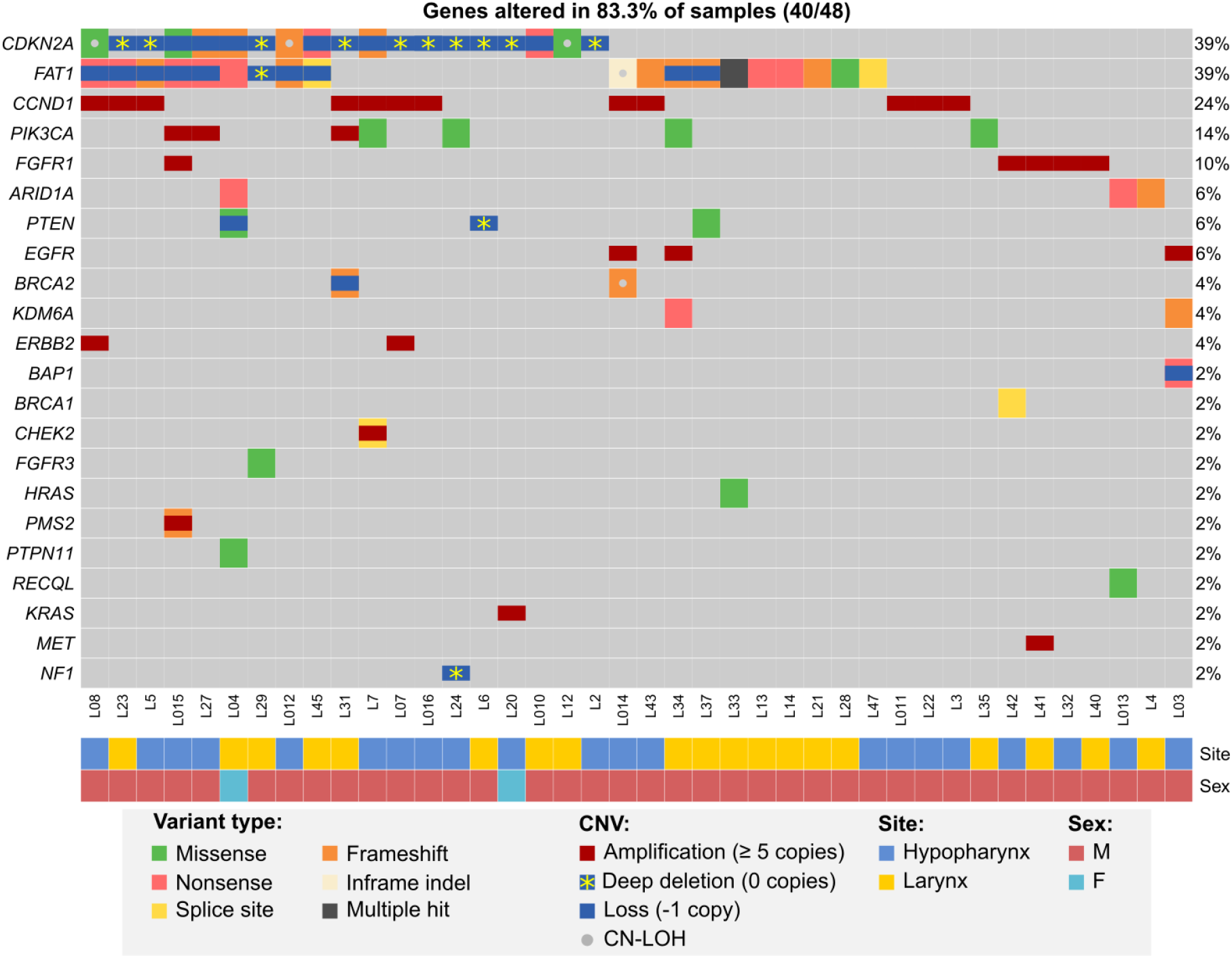
- Potentially actionable or prognostic alterations in hypopharyngeal and laryngeal cancers from the MUW dataset. Included are pathogenic somatic mutation, amplifications (at least 3 additional copies), deep deletions and combinations of mutations and possible second allele losses due to copy loss or CN-LOH.

While we could not reliably assess tumor mutational burden due to limited availability of paired germline samples, we analyzed copy numbers of *CD274/PDCD1LG2* and found that 29.2% (14/48) patients had shallow deletions and 12.5% (6/48) had copy gains (3–4 copies) of these genes, potentially affecting PD-L1 and PD-L2 expression [51] and sensitivity to immune checkpoint inhibitors.

### 3.2. Mutational profile of hypopharyngeal cancer

Finally, to better determine the mutational profile of hypopharyngeal cancers we used data from the MUW dataset as well as previously published data for hypopharyngeal cancer samples with confirmed negative HPV infection status (combined dataset, Supplementary data 3.6). In this analysis, we included 18 significantly mutated genes common to all datasets (listed in supplementary data 1.1). To minimize the differences in variant filtering between datasets we added 6 variants detected in MUW cohort that we excluded from previous analyses due to low likelihood of pathogenicity. In total, 595 non-hypopharyngeal and 67 hypopharyngeal cancers were compared using *maftools*. We did not find any significant differences (Figure 3AB, Supplementary data 2.3) in mutation frequency except for *CASP8* mutations, which were very rare (*p* = 0.0058, OR 0.12, CI 0.0028–0.69) and *HRAS* mutation, which were absent (*p* = 0.025, OR = 0, CI 0–0.87) in hypopharyngeal cancers.

**Figure 3.**
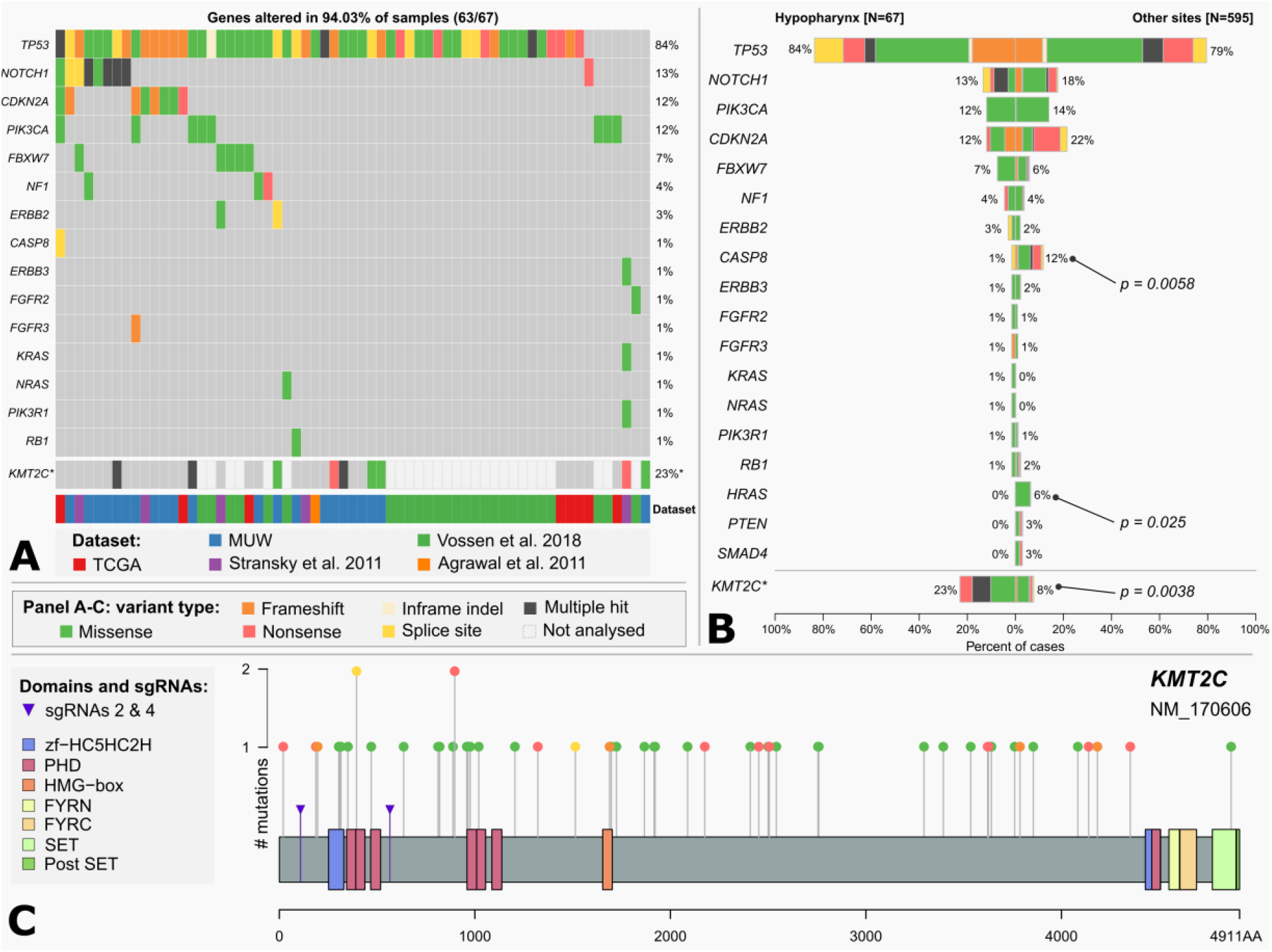
A - Mutational profile of hypopharyngeal cancers. Chart based on a combined dataset and selected genes common to all datasets. B – Comparison of mutation frequency in selected genes between hypopharyngeal and non-hypopharyngeal head and neck cancers. \**KMT2C* mutation frequency is calculated separately excluding NKI dataset (Vossen et al.) in which this gene has not been sequenced. Significance is calculated using maftools mafCompare function (Fisher’s exact test). C – *KMT2C* (MLL3) mutations in head and neck cancers. Additional markers (purple arrows) indicate positions of mutations induced by CRISPR sgRNAs 2 and 4 in FaDu *KMT2C*^mut^ cell lines. Chart is based on a combined dataset except for NKI data.

### 3.3. KMT2C (MLL3) Mutations in Head and Neck Cancer

In our dataset, we identified mutations in *KMT2C* gene, encoding Histone-Lysine N-Methyltransferase 2C, in 14 (29%) or 9 (18.8%) patients, after elimination of benign variants. Seven (14.6%) patients carried *KMT2C* truncating mutations and we also observed frequent shallow deletions or combinations of mutation and CNV/CN-LOH (Figure 1). In the combined HPVneg dataset (excluding NKI dataset, Vossen et al. 2018) *KMT2C* mutations were present in 8.7% (48/551) of patients (Supplementary data 2.3 and 3.7) and 3.4% (19/551) of them harbored truncating mutations that were scattered across the gene (Figure 3C). Moreover, we found that hypopharyngeal cancers harbored more *KMT2C* mutations than cancers from other sites (Figure 3AB, Supplementary data 2.3).

KMT2C was already characterized as a tumor suppressor gene in acute myeloid leukemia. Based on above-mentioned data and our findings in laryngeal and hypopharyngeal cancer, we decided to assess the biological effects of *KMT2C* loss in commercially available HPV-negative hypopharyngeal cell line FaDu (ATCC HTB-43). First, we characterized FaDu cells using targeted NGS and found multiple pathogenic genetic aberrations, including mutations of *TP53* (missense and splice-site), *CDKN2A* (splicesite, homozygous), *FAT1* (frameshift, homozygous), *SYNE1* (missense, frameshift), deep deletions of *AJUBA* and *SMAD4*, high-level amplification of *CCND1*, as well as mutations in *ERBB3, VHL* and other genes (Supplementary data 3.4). Hence, we found that FaDu harbors multiple genetic lesions frequently occurring in head and neck and esophageal cancer.

### 3.4. Induction of KMT2C mutations by CRISPR-Cas9

To study the role of truncating mutations in *KMT2C* in HNSCC, we used CRISPR-Cas9 to induce mutations in the N-terminal quarter of the coding sequence (Figure 3C). We generated *KMT2C*^mut^ FaDu cells stably expressing two different *KMT2C*-targeting sgRNAs (sg2 and sg4) as well as a non-targeting control sgRNA (sgNTC). Using PCR, Sanger sequencing (data not shown) and NGS (Supplementary data 2.4) we confirmed that both sgRNAs effectively induced indels while sgNTC did not, additionally revealing that most clones derived from pools harbored more than 2 copies of *KMT2C*, in concordance with the expected hyperdiploidy of FaDu cells and ploidy analysis in selected cell populations (data not shown). Based on initial growth observations we selected clones sg2-7 and sg4-14 along with clone pools sg2 and sg4 for further experiments.

Targeted NGS analysis of confirmed the specific induction of various *KMT2C* mutations at exons 3 and 12 (Supplementary data 2.4). Furthermore, through Mutect-2 comparisons and manual analysis of NGS data, we did not find any additional, significant genetic point mutations or CNVs unique to any of the modified cell pools, except for clone sg4-14 which could be distinguished by lack of several variants (Supplementary data 3.4) as well as by more pronounced deletion of 3p11.1/*EPHA3*.

### 3.5. The In Vitro Effects of KMT2C Loss on Proliferation and Cisplatin Resistance of Hypopharyngeal Cell Line FaDu

We checked whether *KMT2C* loss affects phenotypic features of FaDu cells and therefore firstly assessed the clonogenic potential of *KMT2C*^mut^ FaDu cells by the clonogenic assay on 6-well plates. We noticed a marked increase in clonogenicity of the modified cells as compared to sgNTC-expressing cells, as manifested not only by significant differences in number of colonies but also in the growth surface area. These effects were confirmed in sg2 and sg4 clone pools (1.19x and 1,47x more colonies and 2.72x and 2.61x larger growth area, respectively) and clones sg2-7 and sg4-14 (1.97x and 2.41x more colonies and 3.34x and 3.44x larger growth area, respectively) (Figure 4AB).

**Figure 4.**
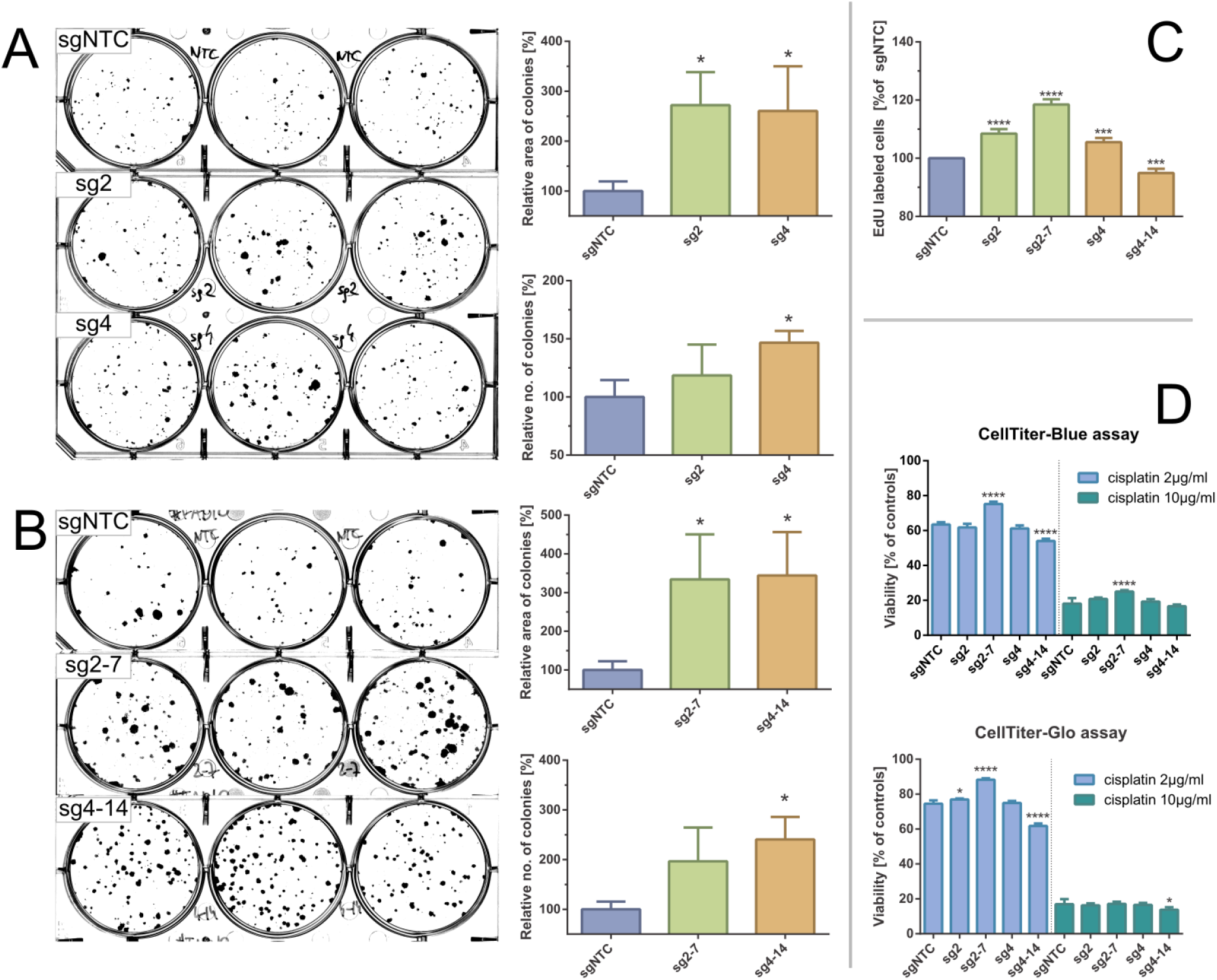
– Effects of CRISPR-Cas9 mediated *KMT2C* mutations on clonogenic potential and cisplating sensitivity of FaDu hypopharyngeal cancer cell line. AB - Clonogenic assays for clone pools (A) and clones (B). Significant increases in colony size and number can be observed in *KMT2C*^mut^ cells as compared to control. C - DNA synthesis / EdU incorporation assay is significantly increased in *KMT2C*^mut^ cells, except for sg4-14 clone. D – Cisplatin sensitivity measured with CellBlue and CellGlo viability assays. Results do not indicate a clear association between *KMT2C* loss and cisplating sensitivity. Control cells (sgNTC) are transduced with non-targeting sgRNA. Asterisks indicate statistical significance, *p-value* in Dunett’s test *\** 0.05 / *** 0.001 / **** 0.0001. Error bars represent standard deviation.

We could not reliably analyze cell cycle using propidium iodide due to variable ploidy among clone pools and clones (data not shown). Instead, we analyzed DNA synthesis rate using EdU incorporation assay. Significantly increased uptake was observed for *KMT2C*^mut^ cells (8.5%, 5.5% and 18.5% increase for sg2, sg4 and sg2-7, respectively), except for sg4-14 clone (5.1% decrease), confirming their proliferative advantage over sgNTC cells (Figure 4C).

Finally, to test if *KMT2C* loss could affect response to standard chemotherapy, we measured viability of FaDu cells exposed for 2 days to two different cisplatin concentrations using CellTiter-Blue and CellTiter-Glo (Promega) assays. While some statistically significant differences in cisplatin sensitivity were observed for clones sg2-7 and sg4-14, they could not be clearly associated with *KMT2C* mutation, given their magnitude and distribution (Figure 4D).

## 4. Discussion

In this work we described the molecular landscape of HPV-negative head and neck tumors located in hypopharyngeal and laryngeal area. In our dataset, we detected molecular alterations, which were previously described in high-throughput head and neck cancer studies [13-15,47,48] as well as new ones. Many of these genetic alterations have well-established or at least putative role in molecular oncogenesis. Importantly, in the majority of analyzed tumors, we found potentially targetable vulnerabilities or prognostic markers.

As expected, the most frequently mutated gene was *TP53*, altered in almost 80% of our samples. Mutations in *TP53* are found in majority of HPVneg head and neck tumors and were shown to affect survival, also in hypopharyngeal cancer; specifically, truncating mutations (frameshift, nonsense, splice-site) seem to be associated with inferior outcome [52,53]. In our study, 45.8% (22/48) patients from the MUW dataset and 43.3% (29/67) of the hypopharyngeal cancer patients in the combined dataset carried truncating *TP53* mutations, possibly leaving others with a more favorable prognosis.

The frequent alterations of cell-cycle genes *CDKN2A* and *CCND1* could predict sensitivity to CDK4/6 inhibition, even if predictive value has not been definitely confirmed yet [54]. Conversely, *FAT1* and *RB1* loss has been shown to negatively affect efficacy of CDK4/6 inhibitors [55]. While 50% (24/48) of our patients harbored mutations in *CDKN2A* and/or *CCND1* (Figure 2), 45.8% (11/24) of them carried concurrent and mostly biallelic *FAT1* alterations that could result in primary resistance. Given the promising clinical activity of palbociclib in HPVneg HNSCC [56], molecular testing for those genes could identify patients with greatest likelihood of response. *FAT1* mutations were also recently associated with progression of HNSCC, which is particularly important given the high mutation rate demonstrated here and in previous studies ([47,57] etc.).

Genes belonging to RTK/RAS/PI3K pathways were affected by evident pathogenic alterations in 43.8% (21/48) of patients (Figure 2) – most of these were RTK amplifications and infrequent hotspot mutations, possibly sensitizing tumors to a wide variety of single-drug or combination therapies. Among other notable genes, *KDM6A* truncating mutations were observed only in two patients (in one of which the mutation was clearly subclonal). Other alterations in this gene included missense mutations and possible deletions on chromosome X. Since *KDM6A* loss has been shown to sensitize cancer cells to EZH2 inhibition (e.g. to FDA-approved tazemetostat) [58] and most of our patients were males, further studies would be desirable. Three patients were found to have truncating *ARID1A* mutations. *ARID1A* mutations negatively impact DNA damage repair in cancer cells and while their role in regulating sensitivity to checkpoint inhibitors is unclear [59,60], they have been also shown to sensitize cancer cells to PARP inhibition [61,62] as well as to EZH2 inhibition [63]. Other genes implicated in DNA damage sensing and repair family were also mutated in our cohort in total of 16% of patients, resulting in a opportunity to use synthetic lethality approach.

Using previously published and new data we further analyze the mutational profile of hypopharyngeal cancer and show that, despite inferior prognosis, it does not differ significantly from other head and neck cancers in terms of mutation frequency in major genes (Figure 3B) which remains in agreement with previous reports [15]. Further analyses on larger cohorts and including CNV data are therefore still required to identify any differences between tumors located in different subsites. Here, we confirmed previous observation that laryngeal and pharyngeal cancers rarely harbor *CASP8* and *HRAS* mutations in comparison to oral cancers [15] – in our comparison, these alterations were almost completely absent in hypopharyngeal tumors when juxtaposed with other head and neck cancer subtypes. Moreover, according to a curated set of non-redundant studies in cBioPortal, *CASP8* and *HRAS* mutations seem to be generally more frequent in head and neck cancers (9.57% and 5.83%, respectively) than in gastroesophageal adenocarcinomas and squamous cell carcinomas (2.07% and 0.15%).

We have detected *pTERT* mutations in 6.4% (3/47) of patients, all originating from larynx (Figure 1). Recent reports indicate that *pTERT* mutations are typically found in oral cancers (up to more than 50%) and rare or absent in other sites [64-66], therefore our results again reaffirm these findings. Consequently, *pTERT* mutations seem to be also very rare in esophageal squamous cell carcinoma [67].

Expression changes and mutations in genes encoding histone methyltransferases have been widely recognized in squamous cell carcinomas [68] and in cancer in general [69]. While the exact nature and roles of these aberrations in specific cancer types are disputable, an extensive experimental evidence suggests that histone methyltransferases frequently serve as tumor suppressors. In our cohort we found *KMT2C, KMT2D* and *NSD1* to be frequently affected by small-scale mutations and copy number alterations (Figure 1). *NSD1-* or histone H3-mutated tumors have been found to constitute a hypomethylated subset within HPVneg HNSCC [70] while *NSD1* and *NSD2* mutations have been associated with favorable prognosis in laryngeal cancers [71]. *KMT2D* mutations have been recently found to sensitize cancer cells to aurora kinase inhibitors in HNSCC [72]. Data on *KMT2C* role in HNSCC is very limited and relatively frequent mutations in our cohort prompted us to conduct additional analyses.

Chen et al. identified *KMT2C* as a target of deleterious mutations as well as copy losses at chromosome 7 and showed its tumor-suppressive role in acute myeloid leukemia, though KMT2C knockdown was incapable to drive oncogenesis alone [73]. In the context of frequent truncating and missense *KMT2C* mutations in breast cancer, Gala et al. found *KMT2C* loss to have both tumor-promoting and tumor-suppressive role depending on the estrogen availability [74]. Cho et al. described frequent *KMT2C* missense mutations in diffuse-type gastric adenocarcinoma that translated into diminished protein expression, which then could be associated with worse prognosis, but only in diffuse-type. In the same study, *KMT2C* loss also induced epithelial-to-mesenchymal transition, including enhanced migration and invasion capabilities [75]. Rampias et al. observed that loss of *KMT2C* activity in bladder cancer and others does not directly affect proliferation or viability but causes DNA repair defects and sensitizes cells to PARP inhibition by downregulation of genes involved in homologous repair of double-strand breaks [76]. This data collectively shows pleiotropic and context-dependent functions of KMT2C and consequences of its aberrations.

KMT2-family genes are altered by various types of mutations in multiple positions. Specifically, *KMT2C* mutations can be located in the SET domain or the PHD domain clusters but are also found in the rest of the gene body and this applies both to missense and truncating variants [69]. As a result, different classes of mutations likely have distinct biological effects e.g., it has been shown that the loss of catalytic activity of *KM2TC* has largely distinct effects from those observed for complete gene inactivation [77,78]. We identified both missense and truncating mutations in *KMT2C*, yet missense mutations were in mostly predicted to be benign or of unknown significance. Moreover, only 6/35 missense *KMT2C* mutation from the combined HNSCC dataset were located in previously described hotspot regions in *KMT2C* [79], being rather scattered across the gene instead. We used CRISPR-Cas9 to induce *KMT2C* inactivation, which should mimic the effects of randomly dispersed truncating mutations described here and by others. This allowed us to find that in cell line FaDu loss of *KMT2C* provides a proliferative advantage, supporting its tumor-suppressive role in hypopharyngeal cancer. It should be emphasized, though, that the effects we measured are possibly limited only to a fraction of cells (Figure 4 AB), which may imply that *KMT2C* disruption is a cooperating event rather than a strong driver of oncogenesis and its outcomes originate from an interplay of many unidentified factors. We also did not obtain a conclusive data linking *KMT2C* loss to cisplatin sensitivity (Figure 4D) which could support previous data on DNA damage repair deficiency [76]. Finally, we found *KMT2C* to be more frequently mutated in cancers originating from hypopharynx than from other sites. However, this difference likely resulted from the limited cohort size as well as inclusion of possibly benign, low-VAF variants in the calculation, that were identified in our samples.

Our study has limitations that warrant further research. We have found limited number of differences in mutation spectrum between cancers located in hypopharynx and others sites, such as larynx, even when combined with previously published data. Additionally, our dataset for hypopharyngeal cancers is limited in respect to cohort size and analyzed genetic alterations. Our data also may be missing some mutations because of the material used for research. Normal DNA samples were available only for 13/49 patients, making it more complicated to discriminate between somatic mutations and rare germline variants. Finally, due to scarcity of reliable hypopharyngeal cancer cell lines, our patient-based data is supported by experiments involving applicable cell line, namely FaDu, though it should be noted that this cell line has a genetic profile typical for this type of cancer.

## 5. Conclusion

In summary, we provide an insight into the molecular landscape of hypopharyngeal cancer and identify new predictive and/or prognostic factors that could improve treatment. Importantly, we confirmed that *KMT2C* plays a tumor suppressor function in hypopharyngeal cancer similarly to hematological malignancies.

## Data Availability

Data presented in this study are available in supplementary materials.

## Supplementary Materials

Supplementary data 1: Supplementary methods, Supplementary data 1.1.: Genes analyzed in MUW and combined datasets, Supplementary data 1.2.: Sequencing panels used to analyze individual samples in the MUW dataset, Supplementary data 1.3.: List of regions commonly covered by all targeted capture probe sets, used in the GISTIC 2.0 analysis of the MUW dataset, Supplementary data 1.4.: Exemplary gating for EdU assays, Supplementary data 2: Supplementary results, Supplementary data 2.1.: All small-scale mutations in SMGs detected in the MUW cohort, Supplementary data 2.2.: GISTIC 2.0 plots generated from MUW and TCGA datasets, Supplementary data 2.3.: Mutations in cancers from hypopharynx vs other sites, Supplementary data 2.4.: *KMT2C* mutations induced by CRISPR-Cas9 in modified cells used in in vitro experiments, Supplementary data 3: Lists of detected mutations, Supplementary data 3.1.: Likely pathogenic variants identified in patients from the MUW dataset, Supplementary data 3.2.: Segmented CNV data for patients from the MUW dataset, Supplementary data 3.3.: Selected genetic alterations of prognostic or predictive significance in the MUW dataset, Supplementary data 3.4.: Selected mutation data for parental FaDu cell line and CRISPR-Cas9 modified clone pools and clones, Supplementary data 3.5.: Segmented CNV data for parental FaDu cell line and CRISPR-Cas9 modified clone pools and clones, Supplementary data 3.6.: All mutations from the combined dataset, Supplementary data 3.7.: *KMT2C* mutations in the combined dataset (excluding NKI dataset).

## Author Contributions

**Conceptualization**, MMM, EOW and TS; **methodology**, MMM, JJ and TS; **software**, MMM and PS; **formal analysis**, MMM; **investigation**, MMM, AR, JJ, MP, KP, AK, MR, JG and MW; **resources**, AR, BG, EOW; **data curation**, MMM, AR, PS, JG and TS; **writing—original draft preparation**, MMM, AR and TS; **writing—review and editing**, JJ, MP, TS; **visualization**, MMM; **supervision**, BG, RP, EOW and TS; **funding acquisition**, MMM and TS. All authors have read and agreed to the published version of the manuscript.

## Institutional Review Board Statement

All patients provided written informed consent to the study, which was approved by our local Institutional Ethics Committee (document number KB/115/2016) and conducted in accordance with the ethical guidelines of the Declaration of Helsinki.

## Informed Consent Statement

Informed consent was obtained from all subjects involved in the study. All patients’ data were kept anonymous.

## Data Availability Statemen

Data presented in this study are available in supplementary materials.

Acknowledgments

Authors would like to thank Marta Kłopotowska, Beata Pyrzyńska, Agnieszka Graczyk-Jarzynka, Małgorzata Bajor, Łukasz Komorowski and Elżbieta Gutowska for excellent scientific and technical assistance.

